# *DIRC3-IGFBP5* is a shared genetic risk locus and therapeutic target for carpal tunnel syndrome and trigger finger

**DOI:** 10.1101/2021.10.07.21264697

**Authors:** Benjamin Patel, Sam O. Kleeman, Drew Neavin, Joseph Powell, Georgios Baskozos, Michael Ng, Waheed-Ul-Rahman Ahmed, David L. Bennett, Annina Schmid, Dominic Furniss, Akira Wiberg

## Abstract

Trigger finger (TF) and carpal tunnel syndrome (CTS) are two common non-traumatic hand disorders that frequently co-occur. By identifying TF and CTS cases in UK Biobank (UKB), we confirmed a highly significant phenotypic association between the diseases. To investigate the genetic basis for this association we performed a genome-wide association study (GWAS) including 2,908 TF cases and 436,579 European controls in UKB, identifying five independent loci. Colocalization with CTS summary statistics identified a co-localized locus at *DIRC3* (lncRNA), which was replicated in FinnGen and fine-mapped to rs62175241. Single-cell and bulk eQTL analysis in fibroblasts from healthy donors (n=79) and tenosynovium samples from CTS patients (n=77) showed that the disease-protective rs62175241 allele was associated with increased *DIRC3* and *IGFBP5* expression. *IGFBP5* is a secreted antagonist of IGF-1 signaling, and elevated IGF-1 levels were associated with CTS and TF in UKB, thereby implicating IGF-1 as a driver of both diseases.

## Introduction

Trigger finger (TF), also known as stenosing flexor tenosynovitis, and carpal tunnel syndrome (CTS) are the two most common non-traumatic hand disorders, with lifetime prevalence of 2-10%^1,2^ and 3-10%^3,4^, respectively. TF causes impaired gliding of the flexor tendons through the first annular (A1) pulley and manifests as painful clicking of the digit during flexion and extension, with progressive stiffness, locking and loss of function. In contrast, CTS is a compression neuropathy of the median nerve that manifests as paresthesia, pain, numbness and weakness of the hand. Both conditions are associated with marked functional impairment and reduced quality of life^5^. CTS is associated with an estimated loss of 78,375 disease adjusted life years annually in the United States, a total disease burden of $2.7-4.8 billion per year^6^ and loss of earnings estimated at $89,000 per patient over 6 years^7^. The mainstay of treatment for both conditions is surgical decompression: release of the A1 pulley in TF; release of the transverse carpal ligament in CTS.

Observational studies have previously suggested a link between CTS and TF: a prospective study found that 43% of patients diagnosed with TF had clinical signs or symptoms of CTS^8^; a study of patients with CTS or TF found that on clinical examination 61% had both CTS and TF^9^. Furthermore, 63% of patients with TF had neurophysiological evidence of CTS compared to 8% of controls^10^. In addition, CTS and TF share risk factors, such as repetitive movements^11^, diabetes^2^ and obesity^12^.

While the genetic architecture of CTS has been investigated recently, through a genome-wide association study (GWAS) in UK Biobank (UKB)^14^, much less is known about the genetic basis of TF. The only published GWAS for TF included only 942 cases and 24,472 controls and identified a single non-replicated locus on chromosome 13^13^. Here, by identifying high-confidence TF cases in the prospectively recruited UKB cohort, we confirm a markedly significant phenotypic association between TF and CTS, and recapitulate known associations between TF and metabolic syndrome. In light of this TF-CTS overlap we hypothesized that there might be a common genetic basis for TF and CTS. To investigate this possibility, we performed GWAS for TF, identifying five independent loci, of which one (*DIRC3*), directly overlapped with the CTS GWAS. Through co-localization analysis and functionally informed fine-mapping, we identify a single putative causal variant at the *DIRC3* locus for both TF and CTS. Finally, using expression quantitative loci (eQTL) analysis in bulk- and single-cell-RNA sequencing datasets, we confirm that this variant modulates expression of the long non-coding RNA (lncRNA) *DIRC3*, and its transcriptional target, insulin-like growth factor-binding protein-5 (*IGFBP5*), directly implementing this pathway in the pathophysiology of TF and CTS.

## Results

### Phenotypic association between CTS and TF

Our phenome-wide analysis re-capitulated a highly significant association between TF and CTS (p<1×10^−300^, odds ratio = 11.97, Figure 1a). To explore the phenotypic association between TF and CTS, we quantified the overlap between TF and CTS (Figure 1b) and explored the clinical characteristics of patients with TF, CTS and TF-CTS overlap (Table 1). We identified evidence for significantly increased prevalence of type 1 and type 2 diabetes mellitus as well as significantly increased HbA1c levels, a routinely-used biomarker for glycemic control, in TF-CTS overlap patients compared to CTS patients (p<1×10^−4^) and TF patients (p<0.01).

**Table 1:** Cohort characteristics, for TF and CTS cases in UKB. P-value refers to Kruskal-Wallis non-parametric test (continuous variable) or chi-squared test for independence (binary variable).

|  | Overlap TF-CTS (N = 887) | Carpal tunnel syndrome only (N = 15,407) | Trigger finger only (N = 2,326) |
| --- | --- | --- | --- |
| **Age of diagnosis** |  |  |  |
| min | 32.72 | 28.74 | 31.66 |
| max | 77.92 | 78.06 | 77.43 |
| mean (sd) | 57.81 ± 8.11 | 57.15 ± 8.92 | 60.09 ± 7.62 |
| **Sex** |  |  |  |
| Male (%) | 272 (31) | 4,634 (30) | 1,003 (43) |
| Female (%) | 615 (69) | 10,773 (70) | 1,323 (57) |
| **BMI** |  |  |  |
| mean (sd) | 29.68 ± 5.59 | 29.48 ± 5.75 | 28.32 ± 4.83 |
| **Height** |  |  |  |
| mean (sd) | 164.12 ± 9.06 | 164.43 ± 8.72 | 166.76 ± 9.14 |
| **HbA1c (IFCC)** |  |  |  |
| mean (sd) | 40.04 ± 11.07 | 37.92 ± 8.44 | 38.75 ± 9.91 |
| **Comorbidities (%)** |  |  |  |
| Current smoker | 94 (11) | 1,728 (11) | 230 (10) |
| Previous smoker | 333 (38) | 5,835 (38) | 959 (41) |
| T1DM | 80 (9) | 524 (3) | 123 (5) |
| T2DM | 191 (22) | 2,476 (16) | 405 (17) |
| Hypertension | 441 (50) | 7,196 (47) | 1,069 (46) |
| High cholesterol | 321 (36) | 5,077 (33) | 888 (38) |
| Gout | 43 (5) | 744 (5) | 118 (5) |
| Osteoarthritis | 463 (52) | 6,749 (44) | 891 (38) |
| **Hypothyroidism** | 65 (7) | 903 (7) | 195 (8) |
| **Rheumatoid arthritis** | 252 (28) | 3,706 (24) | 486 (21) |

**Figure 1:**
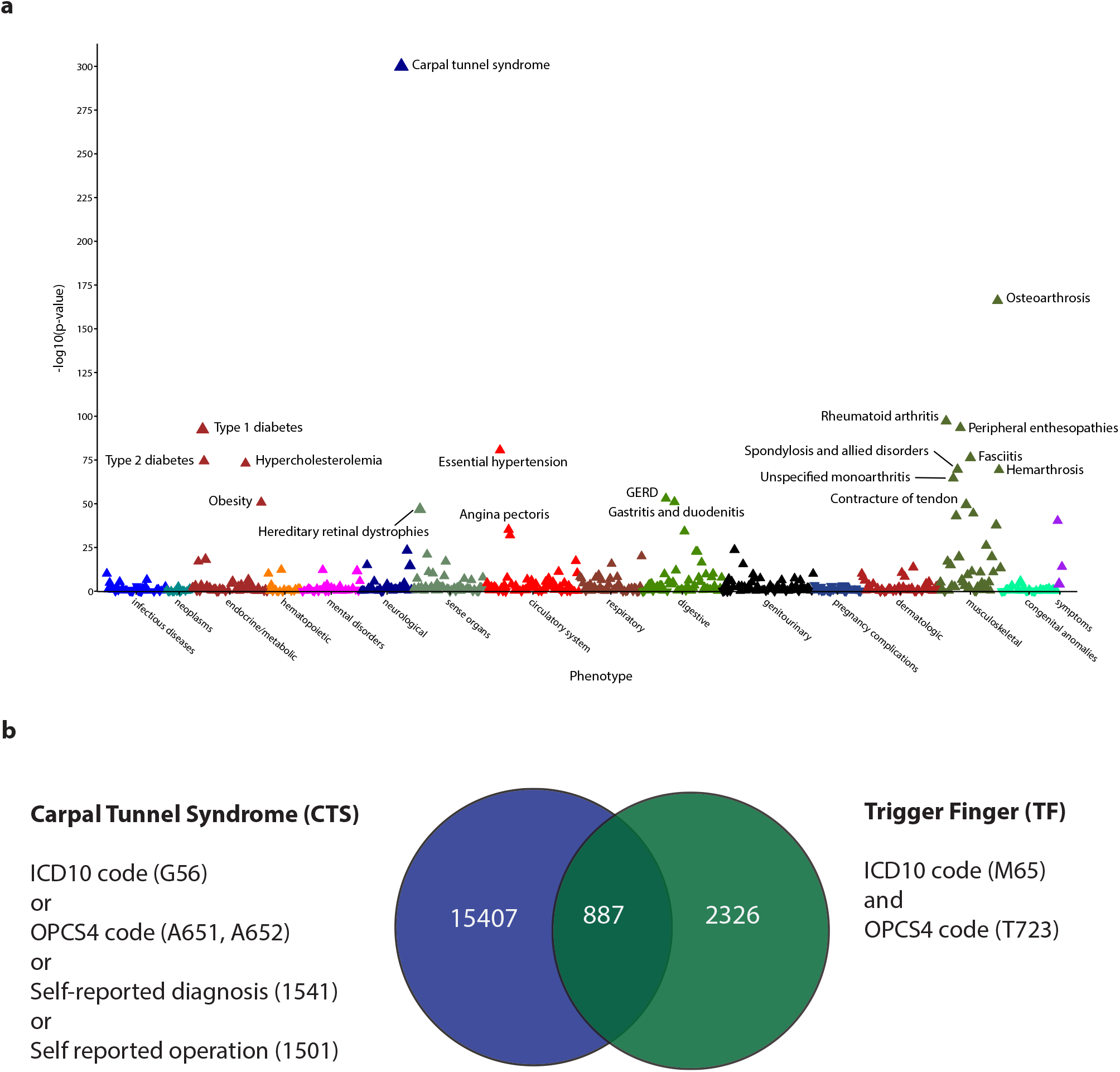
Trigger finger is strongly associated with carpal tunnel syndrome in UK Biobank. (a) Phenome-wide association analysis, showing the association between trigger finger (TF) and 694 phenotypes derived from ICD10 coding in UK Biobank. P-value refers to a Fisher’s exact test, and the direction of the triangle reflects the direction of the effect. (b) Overlap between TF and carpal tunnel syndrome (CTS) in UK Biobank, annotated with the case definition used for CTS and TF cases.

### Genome-wide association analysis for TF

To investigate the genomic architecture of TF, we performed a GWAS in European ancestry UKB participants (n=456,606), incorporating 2,908 TF patients, as previously defined (Figure 1b), and 436,579 controls (Figure S1), using a mixed-model approach, accounting for unbalanced case-control ratios, population structure and cryptic relatedness. There were insufficient TF cases in non-European ancestry groups to perform a sufficiently-powered analysis. There was no evidence of significant confounding with genomic inflation factor (λ_GC_) = 1.058, and we estimated the SNP-based heritability for TF to be 0.8% (SE 0.1%). We identified five independent risk loci comprising 419 variants meeting genome-wide significance (Figure 2a). Conditional analysis revealed no evidence of secondary signals at each locus.

**Figure 2:**
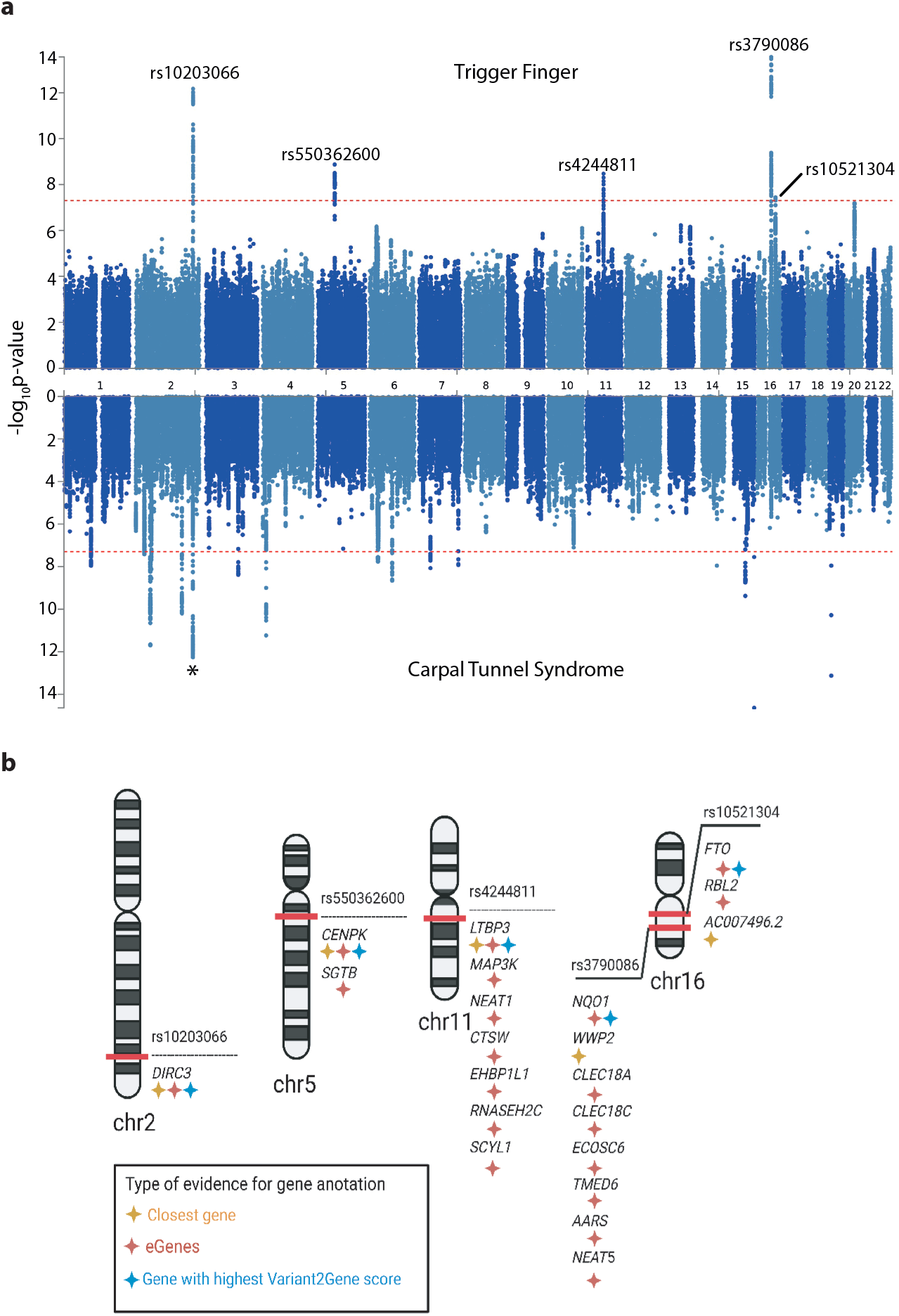
Genome-wide association results for TF and CTS. (a) Summary statistics from TF (upper panel) and CTS (lower panel, derived from Wiberg et al.) genome-wide association analyses in European subjects in UKB. Independent loci in TF summary statistics are annotated with the SNP identifier for the index SNP (lowest p-value) at each locus. Asterisk corresponds to signal at *DIRC3* locus common to TF and CTS analyses. (b) Results of gene prioritization analysis for TF trait, using multiple annotations including nearest gene to SNP, cis-eQTL associations and the Variant2Gene score from OpenTargets Genetics.

**Figure 3:**
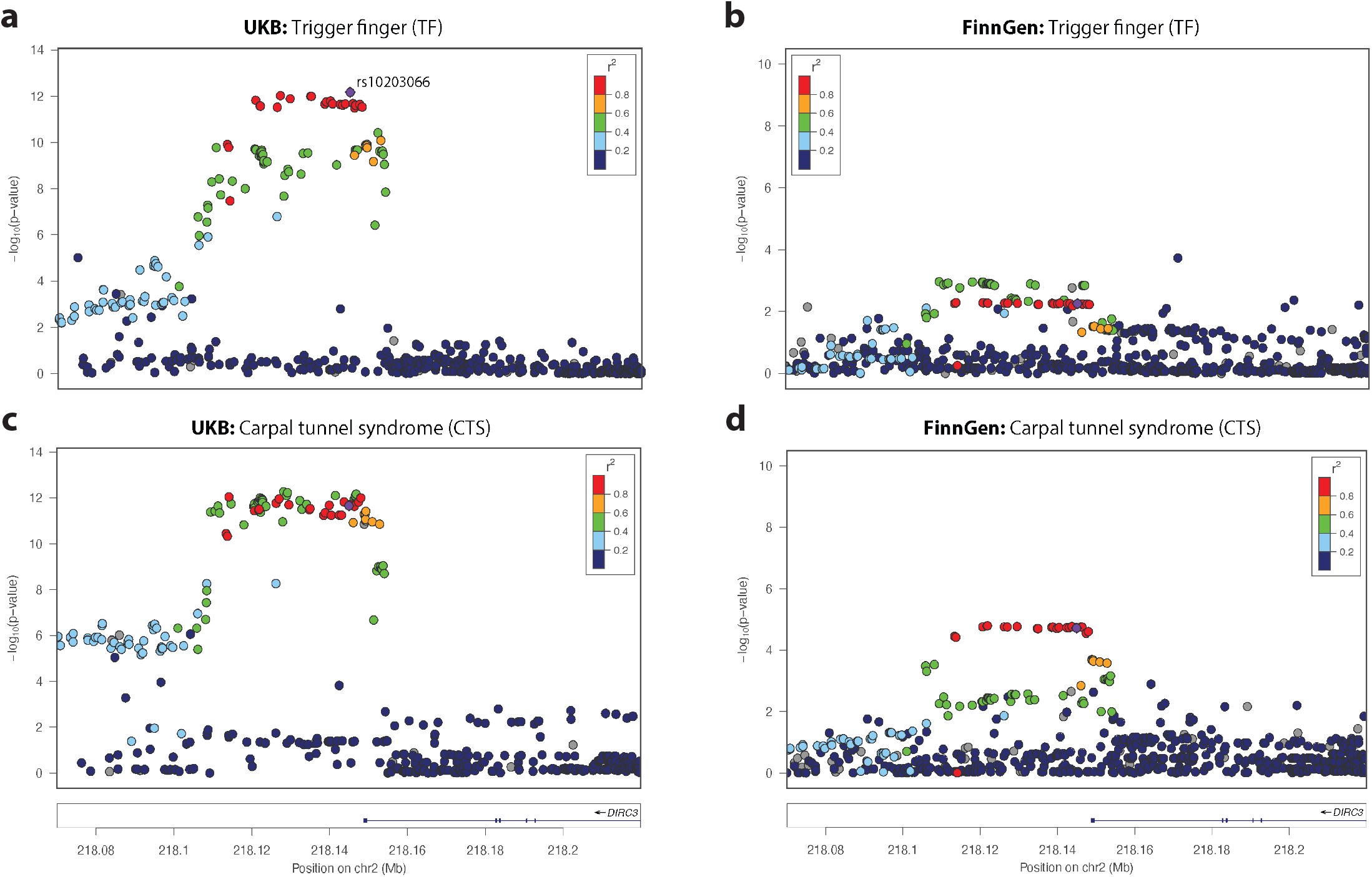
Colocalisation and replication at *DIRC3* locus. SNP-level associations with (a,b) TF and (c,d) CTS associations at *DIRC3* locus, displayed as a LocusZoom plot, derived from (a,c) UKB and (b,d) FinnGen prospectively-recruited cohorts. Index SNP at *DIRC3* locus (rs10203066) is annotated in purple, and each SNP is colored according to r^2^ with rs10203066, derived from 1000 Genomes linkage disequilibrium reference.

Using a multi-modal approach to gene-mapping, we identified 20 candidate genes at the five loci (Figure 2b, summarized in Table 2). In support of our stringent approach to TF phenotype definition, we found that including cases with either ICD-10 or OPCS coding support (extended cohort), or additionally including self-reported TF cases (mixed cohort) markedly reduced the power to detect significance associations across these 5 loci (Table S1).

**Table 2:**
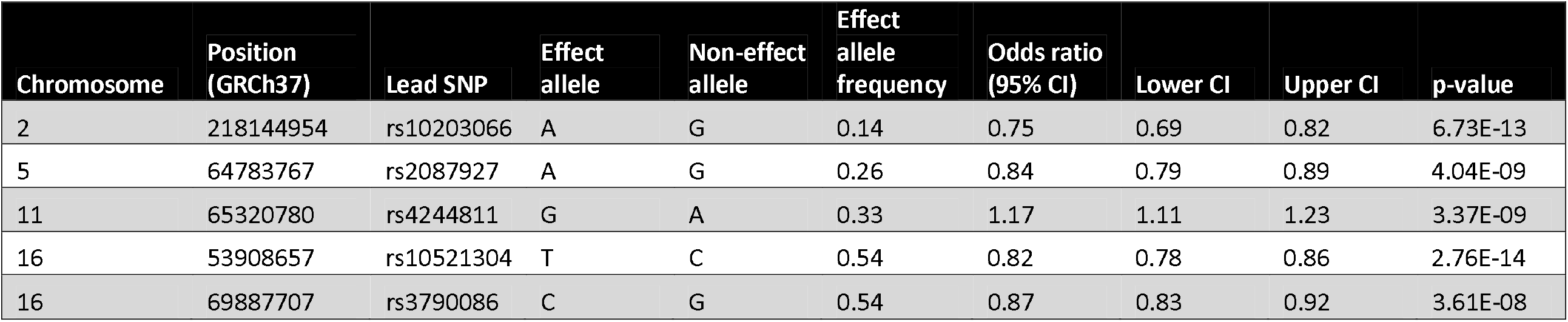
Results of association analysis for TF. Lead SNP refers to SNP with the lowest p-value at each independent locus.

### Co-localization, replication and fine-mapping at *DIRC3* locus

At the *DIRC3* locus, index SNP rs10203066 (p=6.73×10^−13^, OR 0.75, 95% CI 0.69-0.82), was shared with our recently published CTS GWAS (p=2.20×10^−12^, OR 0.88, 95% CI 0.85-0.91). To confirm that this signal was not driven solely by the CTS cases in our TF cohort, we repeated the analysis excluding any TF-CTS overlap cases and confirmed that this signal retains genome-wide significance (p=1.53×10^−10^, Table S1). Consistent with the independent association of this locus with CTS and TF, we confirmed an increase in statistical association having merged TF and CTS cases (p=1.28×10^−24^, Table S1).

To investigate whether this shared signal reflects linkage or a common causal variant, we performed co-localization analysis with a multiple causal variant assumption (SuSiE coloc). This analysis identified a high posterior probability (87%) that TF and CTS share a single causal variant at this locus. To replicate the association between the *DIRC3* locus and both TF and CTS, we extracted summary statistics for TF and CTS from the FinnGen cohort (release 4). The association with both TF (1,485 cases and 137,185 controls; p=0.0055) and CTS (576 cases and 158,705 controls; p=1.90×10^−5^) was confirmed.

We leveraged the co-localization analysis between CTS and TF traits to fine-map this locus by extracting the 95% credible set (n=20) of co-localized variants. Next, using Ensembl Variant Effector Predictor^14^, we functionally annotated the SNPs with their immediate regulatory environment. One SNP, rs62175241, had significant regulatory consequences by disrupting an enhancer site active in fibroblasts as well as the binding motifs for a range of transcription factors including *KLF16* and *KLF18* (Figure S2).

### eQTL analysis at *DIRC3*-*IGFBP5* locus

To investigate how rs62175241 might modulate the expression of the long non-coding RNA (lncRNA), *DIRC3*, we performed expression quantitative trait locus (eQTL) analysis using data from 53 tissues examined as part of the GTEX project^15^. This analysis demonstrated that the effect of rs62175241 (T allele, protective for CTS and TF) on *DIRC3* expression is highly tissue-specific, with positive regulation in stomach and spleen, and negative regulation in testis and amygdala (Figure 4a). In light of evidence that *DIRC3* is able to directly activate expression of *IGFBP5*^16^, with both genes found in the same topologically associating domain (TAD), we examined the effect of rs62175241 on *IGFBP5* expression in GTEx. This again demonstrated tissue-specific eQTL associations and revealed a discordant effect of rs62175241 on *DIRC3* and *IGFBP5* expression in the spleen (Figure 4b). Considering that fibroblast proliferation is a histological feature of both TF and CTS^17^, we further investigated the effect of rs62175241 on *IGFBP5* expression in fibroblasts. We re-analyzed fibroblast single-cell eQTL data from 79 donors^18^. Four of six fibroblast cell sub-types demonstrated a significant positive association between the protective T allele of rs62175241 and *IGFBP5* expression (Table S3), with the strongest association seen in *HOXC6+* cluster (Figure 4c). LncRNAs inherently have markedly lower abundance than mRNAs^19^, and consistent with this and the shallow depth of sequencing in single cell RNA sequencing data, *DIRC3* was detected in less than 1% of cells, precluding further analysis.

**Figure 4:**
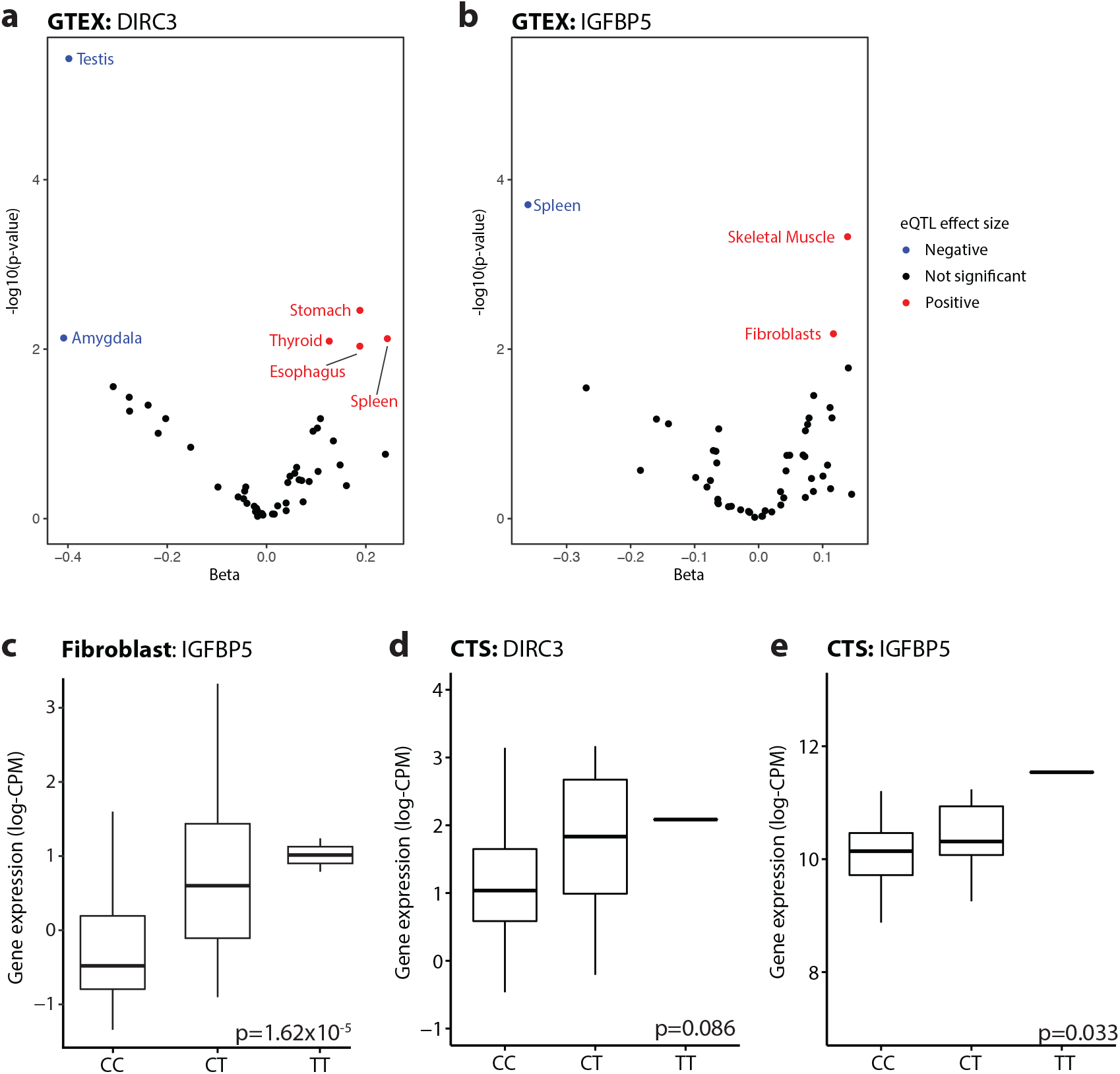
rs62175241 is associated with increased RNA expression the *DIRC3-IGFBP5* axis. Cis-eQTL analysis for rs62175241 on (a) *DIRC3* and (b) *IGFBP5* gene expression in 49 tissues from the GTEx project. Tissue-specific eQTL associations with p-value < 0.01 are annotated, p-value refers to linear model adjusted for PC1-6. (c) Validation of rs62175241 as an eQTL for *IGFPB5* expression in the *HOXC6*+ fibroblast cluster, using single-cell RNA sequencing of fibroblasts from 79 donors. P-value refers to linear model, adjusted for average expression and 1 PEER factor. Association between rs62175241 genotype and (d) *DIRC3* and (e) *IGFBP5* expression derived from paired whole-genome genotyping and RNA sequencing of surgical tenosynovium samples in Oxford-CTS cohort (n=77, n=18 with CT genotype, n=1 with TT genotype). P-value refers to Kruskal-Wallis test.

Next, we analyzed the association of rs62175241 on *DIRC3* and *IGFBP5* expression in diseased tenosynovium samples from patients with CTS (n=77). We confirmed that the protective T allele was associated with significantly increased *IGFBP5* expression (p=0.033, Figure 4d). *DIRC3* expression levels were again low and did not show allele-specific differential expression (p=0.086, Figure 4e). Because *IGFBP5* is a secreted protein, we investigate whether this variant might alter plasma concentration. We analyzed an available plasma proteomic GWAS dataset based on the SomaLogic platform^20^, and found that this variant (tagged by rs10203066, A allele, r^2^=0.99) was associated with a non-significant increased plasma *IGFBP5* (beta=0.017; p=0.11).

### IGF-1 is associated with increased risk of both TF and CTS

As *IGFBP5* is known to directly antagonize IGF-1 signalling^21–23^, with evidence that exogenous growth hormone treatment can cause CTS^21^, we hypothesized that higher IGF-1 plasma levels would be associated with significantly increased risk of both TF and CTS in UK Biobank. We identified significant associations between IGF-1 and TF (hazard ratio (HR) per 1 SD = 1.04, 95% CI 1.01-1.07, p=0.02) and CTS (HR per 1 SD = 1.04, 95% CI 1.02-1.05, p=4.23×10^−6^), which is concordant with a recent CTS-specific analysis in UKB^24^. If the protective effect of rs62175241 was mediated via antagonism of IGF-1 signaling, we hypothesized that this variant would be associated with attenuation of growth hormone-regulated phenotypes such as height and lean body mass. To investigate this, we extracted growth phenotype summary statistics from UK Biobank, and selected traits meeting phenome-wide significance (1×10^−5^). This identified 22 growth-related traits that were significantly associated with rs62175241 (Table S4), all of which had a negative beta, including standing height (p=3.57×10^−18^), weight (p=8.24×10^−6^), forced vital capacity (p=1.13×10^−6^) and appendicular lean mass (p=4.90×10^−33^). Altogether, the available data suggest that IGF-1 is associated with increased risk of TF and CTS, and that the T allele of rs62175241 may act to directly attenuate IGF-1 signaling, thus explaining its protective effect for TF and CTS.

## Discussion

This study confirmed a highly significant phenotypic association between TF and CTS. GWAS identified five risk loci that were significantly associated with TF, one of which was shared with our previous GWAS of CTS patients. Hypothesizing that a single genetic variant might contribute to the pathogenesis of both diseases, we fine-mapped this locus to a single putative causal SNP, rs62175241. Single-cell eQTL analysis demonstrated that this variant was associated with tissue-specific modulation of the expression of *DIRC3* and its known downstream effector, *IGFBP5*. Bulk RNA-seq analysis of surgically resected tenosynovium samples from patients with CTS revealed that the protective variant was associated with enhanced expression of *IGFBP5*. Considering that *IGFBP5* is an antagonist of IGF-1, we found that both TF and CTS were associated with higher levels of IGF-1. These findings are important because they provide direct biological insight into the shared pathophysiological mechanisms contributing to TF and CTS. Furthermore, they provide a starting point for investigating non-surgical interventions for these two common conditions.

The co-localized locus that mapped to the *DIRC3* gene has not been previously described in association with TF. One previous GWAS^13^ has been conducted to identify risk loci associated with TF, finding a single non-replicated locus within *KLHL1* that met significance. *KLHL1* is an actin-binding protein and the authors of this paper speculated that this variant might lead to fibrocartilaginous metaplasia in tenocytes.

In the present TF GWAS, the putative causal SNP, rs62175241, is located 3,731 base pairs from the canonical transcription start site (TSS) of the *DIRC3* gene. The *DIRC3* locus spans 450kb between the *IGFBP5* and TNS1 genes. By mapping the chromatin structure of the *IGFBP5*-*DIRC3*-TNS1 gene territory, Coe et al^16^ have previously identified that *DIRC3* and *IGFBP5* are located within the same topologically-associated domain (TAD) and found two DNA looping interactions between the *DIRC3* locus and *IGFBP5* promoter. *DIRC3* levels positively correlated with *IGFBP5* in melanoma RNA-seq samples and the authors discovered that *DIRC3* acts in cis to control expression of *IGFBP5*.

*IGFBP5* expression appears to be altered in a number of fibrotic disease states. In lung tissue from patients with idiopathic pulmonary fibrosis (IPF), *IGFBP5* was upregulated, and exogenous *IGFBP5* appeared to stimulate ECM secretion by IPF pulmonary fibroblasts^25,26^. Furthermore, *IGFBP5* was upregulated in skin fibroblasts from patients with systemic sclerosis^27^. In the present study, the protective T allele at our putative causal SNP appeared to have tissue-specific effects on expression of *DIRC3* and *IGFBP5* (Figure 4). In both skin fibroblasts and operative tenosynovium samples from patients with CTS, the protective allele was associated with enhanced expression of *IGFBP5. IGFBP5* is a highly conserved, multifunctional secreted protein that binds to IGF and can have complex and varying effects on IGF signaling depending on the tissue type and context^28^. In bone, *IGFBP5* appears to inhibit IGF-1 signalling^29^ by modulating binding to the IGF-1 receptor^22^. Similarly, in mammary tissue, *IGFBP5* appears to regulate involution by inhibiting IGF-1 signalling^23^.

Consistent with the hypothesis that overactive IGF-1 signaling is important in TF and CTS, we discovered that higher circulating IGF-1 plasma levels were significantly associated with increased risk of both conditions. Furthermore, despite CTS generally being associated with decreased height, the protective allele at our putative causal locus was associated with decreased height, potentially suggesting a distinct pathophysiological mechanism. Several other lines of evidence support the role of IGF-1 signaling in TF and CTS. The prevalence of CTS^30^ and TF^31^ is increased in patients with acromegaly, for whom raised IGF-1 levels are characteristic. Furthermore, by normalizing levels of IGF-1, either through pituitary resection or somatostatin analogues, increased tendon thickness at the A1 pulley can be reversed, and symptoms of TF ameliorated^31^. In healthy patients who do not suffer from acromegaly, giving exogenous growth hormone stimulates a rise in IGF-1 and patients subsequently develop CTS^21^. Exogenous growth hormone is also known to increase the risk of type 2 diabetes mellitus, which is significantly enriched in patients with both CTS and TF^32^ (Table 1). Finally, somatostatin analogues work not only by reducing pituitary growth hormone secretion but also by stimulating *IGFBP5* secretion^33^, which in turn inhibits IGF-1 signaling.

We recognize several limitations of the present study. Firstly, our GWAS of TF patients was limited to patients of European ancestry. It would be helpful to further characterize the genomic architecture of TF in non-European groups, who are under-represented in GWAS. While our GWAS has greater power to detect a locus with MAF 50% and OR 1.2 than a previous TF GWAS^13^ (81% versus 3% power), our study was still relatively unpowered, especially for low-frequency variants, meaning that other relevant risk loci may not have met our pre-defined significance threshold. While we were able to replicate our colocalized *DIRC3* locus in both TF and CTS patients in the FinnGen cohort, we were unable to replicate all 5 TF loci. This may be partly explained by the size of the replication cohort (1,485 TF cases), and different case definitions in FinnGen, but may also be consistent with the ‘Winner’s Curse’^34^ phenomenon in GWAS. Regardless, the validation data strongly supports a role for the *DIRC3* locus in both TF and CTS.

We have identified a biologically relevant mechanism that may underpin the association between our co-localized risk locus and TF and CTS. However, we recognize that the association between protective allele, *DIRC3, IGFBP5* and IGF-1 are all correlative and further studies are required to dissect these mechanisms and demonstrate a causative effect.

In conclusion, we have identified a shared variant, associated with both TF and CTS, that partially explains the phenotypic association between these two conditions. Furthermore, we have analyzed a plausible biological model by which this risk is conveyed. Altogether, our findings indicate that the protective variant at rs62175241 acts by enhancing expression of *IGFBP5*, via *DIRC3*, which in turn inhibits IGF-1 signaling. Further studies are required to fully characterize this pathway and delineate whether it might be a valid target for pharmacological management of TF and CTS.

## Methods

### Ethics approval

UK Biobank has approval from the North West Multi-Centre Research Ethics Committee (11/NW/0382), and this study (Genetics and Epidemiology of Common Hand Conditions/GECHCo) has UK Biobank study ID 22572. The Oxford-CTS cohort is derived from two clinical studies approved by the National Research Ethics Service, United Kingdom - Pain in Neuropathy Study (PiNS, 10/H07056/35) and Molecular Genetics of Carpal Tunnel Syndrome (MGCTS, 16/LO/1920).

### UKB genomic data quality control

We selected participants with available imputed genomic data (field 22028), and excluded participants with sex chromosome aneuploidy (field 22019), discordant genetic sex (fields 31 and 22001), excess heterozygosity and missing rate (field 22027). Classification of genetic ancestry was performed as described previously^35^, identifying 456,606 participants with >70% probability of European ancestry. Principal components (PC1-10), accounting for population structure, were computed from high-quality SNPs derived from the ‘in_PCA’ field in the UKB SNP quality control resource (‘ukb_snp_qc.txt’). PCs were computed on the unrelated samples (UKB-provided KING kinship coefficient <0.0442), before being projected onto all European ancestry participants, implemented in the ‘run_pca_with_relateds’ function in the gnomAD package for Hail. For all analyses using genotyped data, we filtered to variants with MAF>1%, call rate>95% and minor allele count>100. For all analyses using imputed data, we filtered to variants with INFO score>0.8 and MAF>1% across the whole cohort.

### Phenotype definition

CTS cases (n=16,294) were defined as previously described^36^ – briefly, we selected individuals with either ICD-10 coding for CTS (G560), OPCS coding for carpal tunnel release (A652), self-reported CTS diagnosis (1541) or self-reported carpal tunnel surgery (1501). To maximize specificity, TF cases were defined by the intersection of patients with ICD-10 coding for TF (M65.3, M65.30-39), and patients with OPCS coding for tendon release (A651, A652). For sensitivity analyses, we explored including patients with either ICD-10 or OPCS coding alone (termed extended cohort) and additionally including patients with self-reported TF symptoms (1619, termed mixed cohort). To explore the common genetic basis of TF and CTS, we defined two additional case groups: 1) TF cases excluding CTS cases and 2) participants diagnosed with either TF or CTS. To define a control group for all subsequent genome-wide analyses, we excluded participants with any of the above diagnosis codes, leaving 482,360 participants, of which 436,576 passed genomic data QC.

### Phenotypic association analysis

To identify diagnoses associated with TF, all UKB ‘first occurrence’ fields and cancer registry data (fields 40005 and 40006) were extracted, with ICD10 codes mapped to Phecodes using established mappings^37^. Data entries were binarized to construct a matrix of 694 diagnosis codes (including TF and CTS, as defined above) versus 502,505 participants in UKB. To determine the association between TF-diagnosis pairs, we constructed a 2×2 contingency table for each pair, and performed a Fisher’s one-way test. Significance level was set at p<1×10^−5^.

### Genome-wide association analysis (GWAS)

Genome-wide association analyses were implemented in Regenie version 2.2.1^38^ in the European ancestry cohort using Firth approximation, with covariates including year of birth (field 34), genotyping array (binarized from field 22000), recruitment center (field 54) and principal components 1-10. For phenome-wide association analysis in UK Biobank, summary statistics were extracted from the OpenTargets Genetics Portal^39^, extracting data specific to the European ancestry group and with p<1×10^−5^.

### Processing of summary statistics

To identify independent signals, we performed conditional and joint analysis, implemented in GCTA-COJO^40^ using a linkage disequilibrium (LD) reference derived from 1000 Genomes European ancestry participants. LD score regression (LDSC) was implemented in the ‘ldsc’ package for R using the UKB European ancestry LD reference derived from the PanUK Biobank project^41^. Lead SNPs were annotated using the OpenTargets Genetics portal^42^ considering three annotations: the nearest coding gene, genes with a cis-eQTL variant in linkage disequilibrium (r^2^>0.8) with the lead SNP, and the Variant2Gene score.

### Co-localization analysis

CTS summary statistics from our previous GWAS were used. SNPs common between the CTS and TF analyses were merged using the ‘snp_match’ function in the ‘bigsnpr’ package for R. To extract data specific to the *DIRC3* locus, we filtered the merged summary statistics to a 1MB region centered at rs10203066. To extract a signed LD correlation matrix for the SNPs in these regions, we used the function ‘ld_matrix_local’ in the ‘ieugwasr’ package for R, using a LD reference derived from 5,000 randomly selected unrelated European participants in UKB. Co-localization analysis was implemented in the ‘coloc’ package for R using the function ‘coloc.susie’, with default parameters. The posterior probability for hypothesis 4 (H4), reflecting existence of a shared causal variant, was extracted. To determine the 95% credible set of colocalized variants, we extracted the posterior probabilities of each SNP, conditioned on H4 being true. These SNPs were functionally annotated using Ensembl Variant Effect Predictor (VEP), release 75 (GRCh37).

### Replication in FinnGen cohort

Summary statistics for TF (M13) and CTS (G6) were downloaded from https://console.cloud.google.com/storage/browser/finngen-public-data-r4/summary_stats/. The FinnGen analysis pipeline has been described previously (https://finngen.gitbook.io/documentation/v/r4/) – briefly, association tests for each endpoint were implemented in SAIGE^43^ adjusted for age, sex, PC1-10 and genotyping batch. LocusZoom plots were generated using http://locuszoom.org/, using legacy mode and the 1000 Genomes European LD reference, relative to the index SNP rs10203066.

### Oxford-CTS cohort sample collection

Sample collection for both the PiNS and MGCTS has been described previously^36^. Briefly, patients with clinically-diagnosed CTS underwent carpal tunnel decompression surgery, during which synovial specimens were collected. Samples were preserved in RNAlater (Thermo Fisher) before extraction using the High Pure RNA Isolation Kit (Roche). For paired whole-genome genotyping, DNA was extracted from whole blood samples using the PureLink Genomic DNA Kit (Invitrogen).

### Oxford-CTS genomic data quality control

For the Oxford-CTS cohort, genotyping was performed using the Infinium Global Screening Array-24 v2.0 (Illumina, San Diego, CA, USA), which includes approximately 700,000 markers. SNPs with call rate <98% or Hardy-Weinberg p<0.0001 were removed, leaving n=691,354 SNPs. Samples with excess heterozygosity (>3 standard deviations from the mean) were removed (n=6), and we confirmed sample call rate >98% and absence of sex discrepancy (using the PLINK --check-sex function). Participants with closer than 3^rd^ degree relatedness (PC-relate kinship coefficient>0.05) were excluded using the pc_relate function in Hail. For classification of genetic ancestry, we used our linkage disequilibrium (LD)-pruned high-quality variants, and merged with variants available from an integrated call-set (call rate>95%) derived from 1000 Genomes and Human Genome Diversity Project (HGDP, gnomAD). European ancestry participants were assigned with a minimum probability of 70%, using a random forest classifier trained using reference data. Bi-allelic SNPs were imputed using the Haplotype Reference Consortium (HRC) reference panel v1.1, with pre-phasing and imputation implemented using EAGLE2 and PBWT respectively (Sanger Imputation Server).

### RNA sequencing (RNA-seq)

RNA extraction and library preparation were performed as described previously^36^. Reads were aligned to GRCh37 reference with STAR^44^ using the Ensembl 87 gene annotation, with gene-level counts assigned using HTSeq^45^. Count-level batch correction between GECHCo and PiNS cohorts was performed using ComBat-seq. In order to facilitate inter-sample comparisons, count-level data was TMM-normalized and log-transformed to generate log-transcripts per million (log-CPM) data.

### eQTL analysis

Harmonized summary statistics from analysis of Genotype-Tissue Expression (GTEx) project data were downloaded from eQTL Catalogue^15^. eQTL analysis for *IGFBP5* in the Neavin et al. cohort was performed as described previously^18^. Briefly, for each individual and fibroblast cluster, the quantile-normalized pseudobulk average expression was extracted, and cis-eQTL association statistics were computed using a linear model implemented in MatrixEQTL^46^, with one PEER factor as covariate. eQTL analysis for *DIRC3* and *IGFBP5* in the Oxford-CTS cohort was implemented as a Kruskal-Wallis test for gene expression (log-CPM) against genotype.

### IGF-1 in UK Biobank

IGF-1 plasma levels from the first recruitment visit were extracted from field 30770, and were normalized by Z-scoring, stratified by age (decile) and sex. For Cox regression of TF-or CTS-free survival against Z-scored IGF-1, the time variable used was time from blood sampling to ICD-coded diagnosis of CTS or TF, or last follow-up date. Last follow-up date was determined through integration of death status, recruitment visits and ICD coding dates. The model was adjusted for age at first recruitment visit, sex and recruitment center.

## Data Availability

RNA sequencing data from the PiNS cohort has been reported previously and is available at accession GEO108023. The expanded Oxford-CTS RNA sequencing cohort, as reported in this manuscript, will also be made available through SRA and GEO. Normalized batch-corrected gene expression data with matched genotypes at rs62175241 will be provided as source data. Single-cell fibroblast eQTL summary statistics are published alongside the original manuscript18. UK Biobank data can be requested through the application process detailed at https://www.ukbiobank.ac.uk/. Notebooks detailing the core genomic analyses performed (cohort quality control and GWAS) will be made available on Github. Summary statistics for GWAS will be uploaded to GWAS Catalog and the Oxford Research Archive.

## Author contributions

B.P, D.F. and A.W. conceived and designed the study. B.P. constructed statistical analysis plan and extracted case and control cohorts. S.K. performed computational analyses. G.B. assimilated and processed raw RNA-seq data. W.A. extracted DNA for genotyping, and M.N. performed quality control and imputation of the genotype data. A.B.S. and D.B. recruited and phenotyped the PiNS cohort, and performed RNA extraction with A.W. D.N. completed single-cell analyses, with supervision from J.P. D.F. and A.W. supervised the project and guided all data analysis. B.P., D.F. and A.W. wrote the manuscript with input from all authors. All co-authors approved the final version of the manuscript.

## Competing interests

All authors declare no conflicts of interest.

## Data and code availability

RNA sequencing data from the PiNS cohort has been reported previously and is available at accession GEO108023. The expanded Oxford-CTS RNA sequencing cohort, as reported in this manuscript, will also be made available through SRA and GEO. Normalized batch-corrected gene expression data with matched genotypes at rs62175241 will be provided as source data. Single-cell fibroblast eQTL summary statistics are published alongside the original manuscript^18^. UK Biobank data can be requested through the application process detailed at https://www.ukbiobank.ac.uk/. Notebooks detailing the core genomic analyses performed (cohort quality control and GWAS) will be made available on Github. Summary statistics for GWAS will be uploaded to GWAS Catalog and the Oxford Research Archive.

## Acknowledgments

This work was conducted using the UK Biobank resource under application number 22572. We thank all patients and their families who have volunteered to participate in clinical research. We thank the Oxford Genomics Centre at the Wellcome Centre for Human Genetics (funded by Wellcome Trust grant reference 203141/Z/16/Z) for the generation and initial processing of RNA sequencing data. We also thank our clinical collaborators of the NDOMRS Hand Research Group for the collection of surgical specimens (https://www.ndorms.ox.ac.uk/research-groups/collaborative-hand-research-group). S.O.K. is supported by the Starr Centennial Scholarship at the Cold Spring Harbor Laboratory School of Biological Sciences. Computational analyses at Cold Spring Harbor Laboratory were performed with assistance from the US National Institutes of Health Grant S10OD028632-01. A.W. is supported by a NIHR Clinical Lectureship and a grant from the MRC DTP (MR/N013468/1). D.F. is supported by the NIHR Biomedical Research Centre, Oxford (BRC). A.S. is supported by a Wellcome Trust Clinical Career Development Fellowship (222101/Z/20/Z) and the National Institute for Health Research (NIHR) Oxford Biomedical Research Centre (BRC). The views expressed are those of the authors and not necessarily those of the NHS, the NIHR or the Department of Health.

---

**Figure S1:**
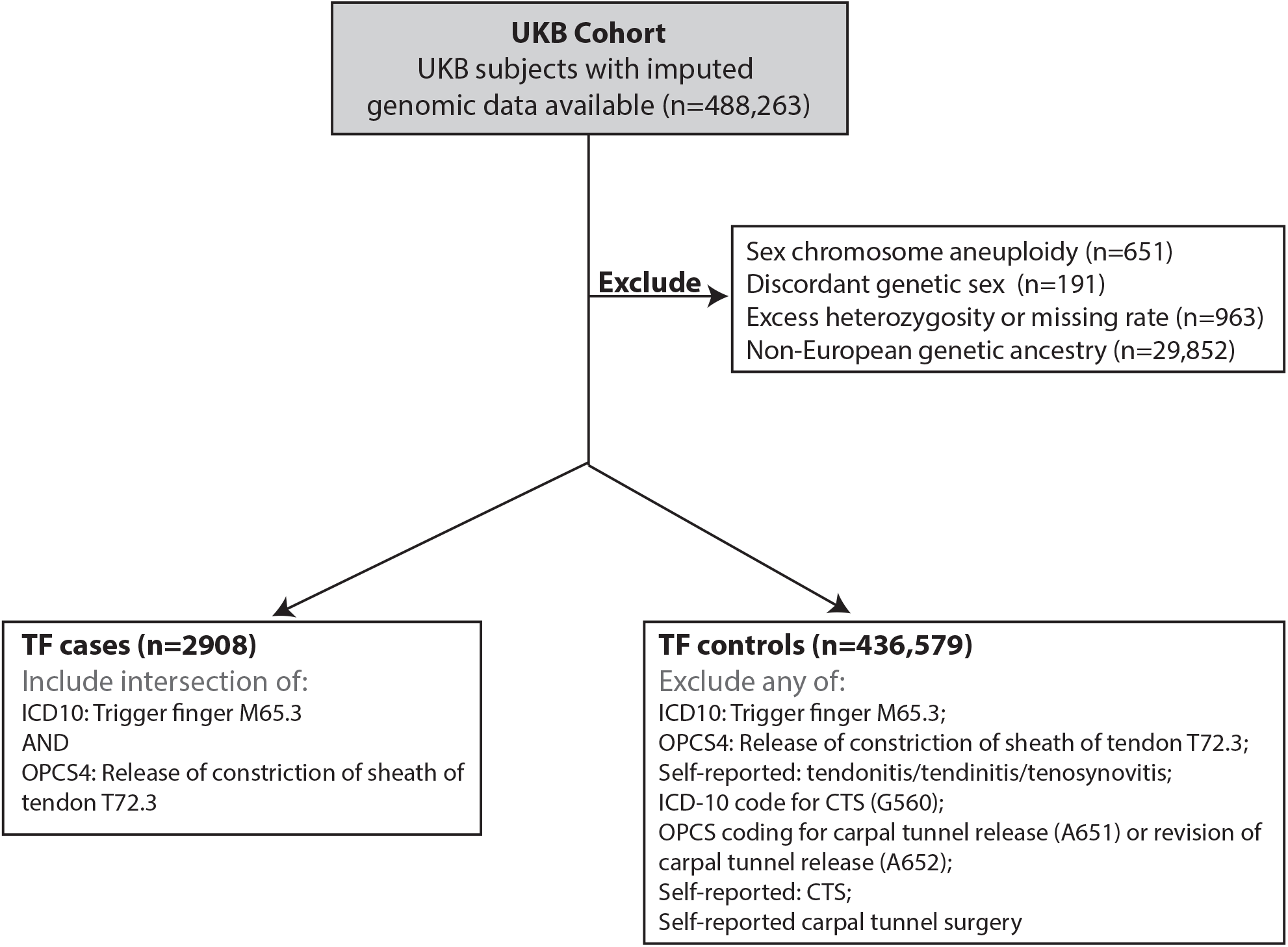
CONSORT diagram summarizing the selection of case (n=2908) and control (n=436,579) cohorts for trigger finger (TF) genome-wide association analysis in European subjects from UKB cohort.

**Figure S2:**
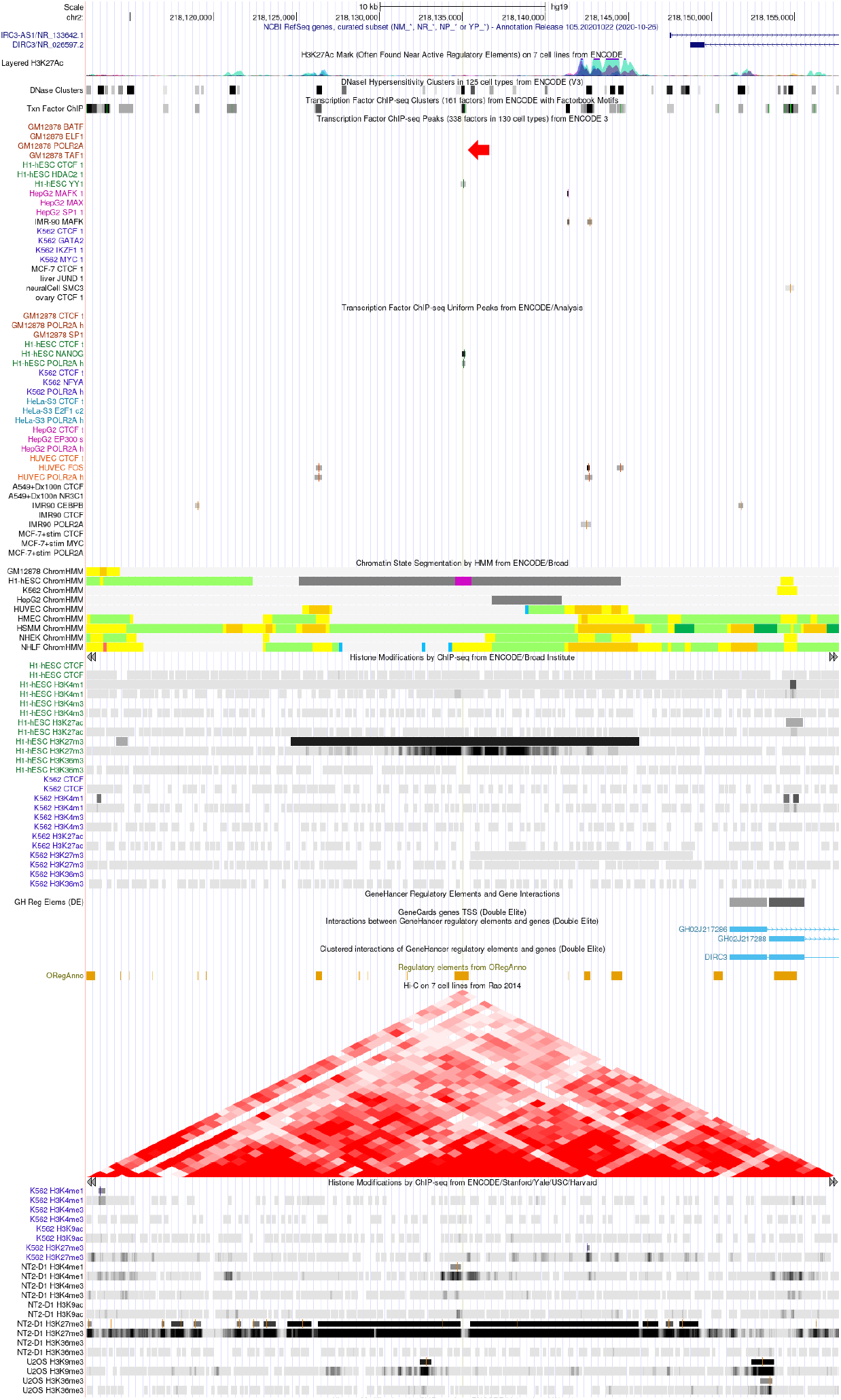
Genomic context of rs62175241 (marked with arrow), annotated with *DIRC3* transcript location, functional genomics annotations including transcription factor and histone modification ChIP-seq, DNase-accessible sites, predicted regulatory elements and Hi-C data indicating chromosome spatial organization. Credit UCSC Genome Browser (https://genome.ucsc.edu/index.html).

## Notes

### Competing Interest Statement

The authors have declared no competing interest.

